# CC180 clade dynamics do not universally explain *Streptococcus pneumoniae* serotype 3 persistence post-vaccine: a global comparative population genomics study

**DOI:** 10.1101/2024.08.29.24312665

**Authors:** Sydney Stanley, Catarina Silva-Costa, Joana Gomes-Silva, Jose Melo-Cristino, Richard Malley, Mario Ramirez

**Author notes:** Correspondence to: Dr. Sydney Stanley, Boston Children’s Hospital and Harvard Medical School, Boston, MA, USA. Co-senior authors (Richard Malley, Mario Ramirez). Contributions: Conceptualization: SS, RM, MR; Methodology: SS, MR; Software: SS; Formal analysis: SS, CSC; Investigation: CSC, JGS, JMC; Resources: RM, MR; Data curation: CSC, JMC; Writing – original draft: SS; Writing – review & editing: SS, CSC, JGS, JMC, RM, MR; Visualization: SS; Supervision: RM, MR; Funding acquisition: JMC, RM, MR.

## Abstract

**Background:** Clonal complex 180 (CC180) is currently the major clone of serotype 3 *Streptococcus pneumoniae* (Spn). The 13-valent pneumococcal conjugate vaccine (PCV13) does not have significant efficacy against serotype 3 despite polysaccharide inclusion in the vaccine. It was hypothesized that PCV13 may effectively control Clade I of CC180 but that Clades III and IV are resistant, provoking a population shift that enables serotype 3 persistence. This has been observed in the United States, England, and Wales but not Spain. We tested this hypothesis further utilizing a dataset from Portugal.

**Methods:** We whole-genome sequenced (WGS) 501 serotype 3 strains from Portugal isolated from patients with pneumococcal infections between 1999-2020. The draft genomes underwent phylogenetic analyses, pangenome profiling, and a genome-wide association study (GWAS). We also completed antibiotic susceptibility testing and compiled over 2,600 serotype 3 multilocus sequence type 180 (MLST180) WGSs to perform global comparative genomics.

**Findings:** CC180 Clades I, II, III, IV, and VI distributions were similar when comparing non-invasive pneumonia isolates and invasive disease isolates (Fisher’s exact test, P=0.29), and adult and pediatric cases (Fisher’s exact test, P=0.074). The serotype 3 CCs shifted post-PCV13 (Fisher’s exact test, P<0.0001) and Clade I became dominant. Clade I is largely antibiotic-sensitive and carries the ΦOXC141 prophage but the pangenome is heterogenous. Strains from Portugal and Spain, where Clade I remains dominant post-PCV13, have larger pangenomes and are associated with the presence of two genes encoding hypothetical proteins.

**Interpretation:** Clade I became dominant in Portugal post-PCV13, despite the burden of the prophage and antibiotic sensitivity. The accessory genome content may mitigate these fitness costs. Regional differences in Clade I prevalence and pangenome heterogeneity suggest that clade dynamics is not a generalizable approach to understanding serotype 3 vaccine escape.

**Funding:** National Institute of Child Health and Human Development, Pfizer, and Merck Sharp & Dohme.

**Research in context:** *Evidence before this study:* We conducted this study because of the mounting interest surrounding the changing prevalence of serotype 3 *Streptococcus pneumoniae* (Spn) genetic lineages and the potential association with escape from 13-valent pneumococcal conjugate vaccine (PCV13) control. To inform our investigation, we searched the PubMed database using different combinations of the following keywords: “Streptococcus pneumoniae”, “serotype 3”, “CC180”, “PCV13”, “Clade Iα”, “Clade Iβ”, and “Clade II”. The search included all English language primary research articles published before July 1^st^, 2024; this language limitation may bias the results of our assessment. Most ST3 isolates belong to clonal complex 180 (CC180), and one study identified three major lineages within CC180: Clade Iα, Clade Iβ, and Clade II. This study observed a global trend of increasing Clade II prevalence with a concomitant decrease in Clade I prevalence over time, which was associated with the introduction of PCV13 in the United States. A report from England and Wales made a similar observation. It was therefore hypothesized that PCV13 may be effective at controlling Clade Iα and that Clade II is driving vaccine escape. Later work refined the clade classification system as follows: Clade I (Clade Iα), Clades II and VI (Clade Iβ), Clades III and IV (Clade II), and Clade V. Clade I strains are marked by a significantly lower recombination rate partly due to the presence of a lineage-specific prophage interfering with competence development, which is a potential mechanism explaining the possible reduced fitness of Clade I. Clade I is also noted to be mostly antibiotic-susceptible. However, a recent study found that Clade I persists as a dominant serotype 3 lineage in Spain, so the generalizability and implications of clade dynamics remain unclear.

*Added value of this study:* Early work assessing the association between changes in serotype 3 clade prevalence and PCV13 was limited by small sample sizes. In addition, studies investigating differences in clade dynamics did not comprehensively consider patient age or disease manifestations such as non-invasive pneumonia and invasive infections. In this study, we evaluated 501 serotype 3 strains from Portugal to investigate clade dynamics. This must be explored in different geographic contexts for a more robust understanding of changing serotype 3 population genomics. We also sought to define genetic determinants linked to strains from regions in which Clade I remains dominant. This is an important step towards a more mechanistic understanding of the serotype 3 CC180 lineage fitness landscape.

*Implications of all the available evidence:* Unlike other serotypes covered by PCV13, serotype 3 has evaded vaccine control. It has been suggested that Clade I prevalence has decreased due to PCV13, which has created an expanded niche for strains from other clades and ultimately renders PCV13 less effective against serotype 3. This postulation has important implications for the future design of an improved vaccine, so this hypothesis must be thoroughly tested in diverse contexts. We find that Clade I remains the dominant lineage in Portugal even after the introduction of PCV13. We delineate Clade I pangenome heterogeneity and show that strains from Portugal and Spain share similar pangenome features in contrast to Clade I strains from regions where Clade I decreased in prevalence, which should motivate future studies to elucidate more generalizable population genomics trends that may better inform strategies for the design of an improved vaccine.

## Introduction

*Streptococcus pneumoniae* (Spn) is a major threat to global health as the bacterium is the primary causative agent of lower respiratory infections that result in significant morbidity and mortality, especially in young children and older adults (1). Spn infection can present as pneumonia, mild cases of sinusitis and otitis media, or severe invasive pneumococcal disease (IPD) such as bacteremia or meningitis. In all, Spn is responsible for over 300,000 deaths in children worldwide and IPD is associated with up to 20% case-fatality in adults (2,3).

The 13-valent pneumococcal conjugate vaccine (PCV13) targets the antigenic capsular polysaccharide (CPS) of 13 different Spn serotypes that are associated with IPD. National childhood PCV13 programs in several countries have resulted in decreased incidence of both pediatric and adult IPD caused by most serotypes included in the vaccine (vaccine types or VTs), except for serotype 3 (4-7). This includes Portugal, where PCV13 first became widely available in the private market in 2010 and later included as part of the national immunization program in 2015 with >95% uptake; the overall proportion of IPD caused by PCV13 VTs dropped significantly but serotype 3 remained a main contributor (8-10). Further, serotype 3 is the main cause of adult pneumonia in Portugal (9). As for children, serotype 3 has become the leading cause of pediatric complicated pneumococcal pneumonia, which involves pneumonia along with pleural effusion or empyema (11).

It is unclear why serotype 3 has a high propensity for severe disease manifestations and has resisted PCV13 control. Serotype 3 has a unique CPS production mechanism that enables capsular shedding, which may contribute to these processes (12). To better understand the association between serotype 3 and PCV13 more specifically, previous studies have evaluated the impact of vaccination on bacterial population genomics. Clonal complex 180 (CC180) is the primary circulating clone of serotype 3. Azarian et al. reported that CC180 is comprised of three major lineages: Clade Iα, Iβ, and II (13). They also observed a global trend where Clade II replaced Clade Iα as the most prevalent lineage over time (13). Notably, this shift was associated with the introduction of PCV13 in the United States but they could not show this in other regions, potentially due to small sample sizes (13). Consistent with this observation, Clade II has also become the most common lineage in England and Wales (14,15). The clades were later subdivided as follows to acknowledge additional sublineages of interest: Clade I (Clade Iα), Clades II and VI (Clade Iβ), Clades III and IV (Clade II), and Clade V (16). It is unclear why Clade I is being outcompeted by other lineages. Fitness may be hampered by a low recombination rate, mediated by the insertion of the ΦOXC141 prophage, and general antibiotic susceptibility (13,16). However, a recent report from Spain observed that Clade I remained the dominant lineage post-PCV13, so the generalizability of impaired Clade I fitness and the putative link between PCV13 and the changing serotype 3 population structure remain unknown. This is important to assess as the field seeks potential avenues to optimize vaccine efficacy against serotype 3. In addition, serotype 3 clades have not been comprehensively considered in relation to pediatric vs adult cases and non-invasive pneumonia (NIP) vs IPD. Towards this end, we investigated the clade dynamic of 501 clinical serotype 3 strains from Portugal. We assessed the association between clade background and patient age, disease presentation, and the introduction of PCV13. We also compiled a global dataset of over 2,600 serotype 3 strains to identify genomic and genetic commonalities underlying country-level clade epidemiological patterns.

## Methods

### Study design, participants, and Spn isolates

Between 1999-2020, we received 14,506 Spn isolates recovered from human infections in hospitals throughout Portugal for which we determined the serotype. These include all isolates recovered from invasive infections, i.e. in which Spn were recovered from normally sterile samples (blood, cerebrospinal fluid, pus from otomastoiditis or other deep tissue collections, synovial fluid, peritoneal fluid, pericardial fluid and pleural effusion), and a sample of isolates recovered from NIP, i.e. in which Spn were recovered from respiratory tract products from patients with a presumptive diagnosis of pneumonia (sputum, bronchial secretions and bronchoalveolar lavage). Of these, 2,551 isolates were recovered from the pediatric age group (<18 years) and 11,953 were recovered from adults (>18 years). A total of 8,850 corresponded to invasive isolates (yearly average number 402, range 132-517) and 5,656 to NIP isolates (yearly average number 257, range 629-87). The proportion of serotype 3 Spn presented yearly fluctuations and tended to be higher in adults (invasive isolates yearly average 13.2%, range 8.5%-17.1%; NIP isolates yearly average 16.7%, range 9.2%-28.3%) than in the pediatric population (invasive isolates yearly average 6.0%, range 0%-20.8%; NIP isolates yearly average 7.8%, range 0.0%-17.0%). We selected for genomic analysis 350 serotype 3 invasive isolates (304 from adults and 46 from pediatric patients, corresponding to 29% and 64% of invasive serotype 3 isolates from these age groups, respectively) and 147 serotype 3 NIP (123 from adults and 24 from pediatric patients, corresponding to 18% and 25% of NIP serotype 3 isolates from these age groups, respectively). We also included four samples that were not classified in any of these two infection types. In addition, not all years were sampled. Invasive isolates were selected from all years (1999-2020) but the NIP isolates from adults were selected only from 2016-2020 (yearly average 46.1% of available isolates) and from pediatric patients from 2014-2020 (yearly average 59.5% of available isolates). Given the introduction of PCV13 in the private market in Portugal in 2010, we considered a pre-PCV13 period (1999-2010) and post-PCV13 period (2011-2020) in our analyses.

No personal and sensitive data was collected. The data collected is anonymous data, i.e., the identity of the person to whom the data are referred to was unknown. The patient samples are collected within the context of the diagnostic workup at the discretion of the attending physician and no specific guidelines or recommendations are in force because of the study. The study was approved by the Institutional Review Board of the Centro Académico de Medicina de Lisboa (240/22).

### Pneumococcal growth conditions, antibiotic susceptibility testing and genomic sequencing

Spn were grown in blood agar plates or brain heart infusion (BHI) broth (Oxoid, Basingstoke, UK) as appropriate. The minimum inhibitory concentrations (MICs) of penicillin and cefotaxime were determined by Etest (Biomérieux, Marcy L’Étoile, France). We obtained penicillin MICs for 401 of the 501 strains (80%) and cefotaxime MICs for 387 strains (77%). Susceptibility to cotrimoxazole, erythromycin, clindamycin, tetracycline, chloramphenicol, levofloxacin, linezolid, and vancomycin was evaluated via disk diffusion testing (Oxoid, Basingstoke, UK). Antibiotic susceptibility data is available for 393 of the 501 strains (78%) except for linezolid and vancomycin (385 strains or 77%). Antimicrobial susceptibility was determined according to the European Committee on Antimicrobial Susceptibility Testing guidelines for all antibiotics except for chloramphenicol, for which we utilized the Clinical and Laboratory Standards Institute guidelines (17,18).

Genomic DNA was extracted and purified from overnight cultures in BHI broth with the PureLink™ genomic DNA minikit (Invitrogen, Carlsbad, CA, USA), adding deoxycholate to the bacterial lysis step. Libraries were prepared with the Nextera DNA library preparation kit (Illumina, San Diego, CA, USA) and sequenced with an Illumina MiSeq or NextSeq instrument.

### Phylogenetics

We prepared the paired-end FASTQ files generated from the Illumina sequencing for downstream processing with the Sickle tool (version 1.33) to trim reads with a quality threshold of 20 and length cutoff of 30. TrimGalore (version 0.6.6) was used for further quality and adapter trimming (19). Reference-based genome assembly was performed with Snippy (version 4.6.0) using Spn OXC141 (NCBI accession: NC_017592), a serotype 3 multilocus sequence type 180 (MLST180) strain belonging to Clade I (13). Core genome single nucleotide polymorphism (SNP) phylogenetic trees with recombination removed were generated via Gubbins (version 3.3.1) with the IQ-TREE setting; we used FastTree (version 2.1.10) to provide a starting tree (20-22). Trees were visualized with iTOL (23). A serotype 11D strain (NCBI accession: ERR3343678) was used as the outgroup for the phylogenetic tree with the 501 serotype 3 strains from Portugal but the outgroup was removed from the final tree. The phylogenetic tree with the 2,663 serotype 3 MLST180 isolates, including those from Portugal, comprised WGSs sourced from the NCBI database and PubMLST (24). We confined this dataset to MLST180 because previous publications exploring CC180 lineage dynamics overwhelmingly belong to this sequence type. A serotype 3 strain belonging to MLST4735 (NCBI accession: ERR461187) was used as the outgroup for this tree.

### MLST, Clonal complex, Global Pneumococcal Sequence Clusters (GPSCs), and clade identification

Strain MLST was determined with the MLST tool (version 2.19.0) (24). This required *de novo* genome assembly of the cleaned reads, which was completed with Spades (version 4.0.0). Clonal complex (CC) was determined by identifying single-locus variants (SLVs), they were named according to the most common ST within a given CC. The CC plot was created with GrapeTree (version 1.5.0) (25). GPSCs were identified with popPUNK (26) and clades were identified in reference to those described by Azarian et al. and Kwun et al. (13,16).

### Resistance genes categorization

Antibiotic resistance genes were characterized from the cleaned reads with ARIBA (version 2.4.16) using the ARG-ANNOT and CARD databases (27-29). We employed pbptyper (version 1.0.4) to classify penicillin-binding protein gene alleles from contigs (30).

### Phage annotation

We screened the assembled contigs for the ΦOXC141 prophage (NCBI accession: KY065494) with the PhaBOX pipeline (31). We denoted strains as phage positive if a contig mapped to at least 95% of the ΦOXC141 sequence.

### Pangenome comparisons

Contigs were annotated with Prokka (version 1.14.6) and polished with Panaroo (version 1.5.0) using the option to filter out putative pseudogenes, gene fragments, and genes of abnormal lengths (32-33). We created a t-distributed stochastic neighbor embedding (tsne) plot to cluster the pangenome content with the Rtsne (version 0.17) R (version 4.4.0) package. We also performed a genome-wide association study (GWAS) with Pyseer to identify genes associated with countries where Clade I remains dominant (34). We utilized the linear mixed model option, the gene presence and absence output from Panaroo, and controlled for population structure by constructing a phylogeny-based similarity matrix. Strain country of origin was classified according to the following publications: Portugal (this study), United States (13), England and Wales (14), and Spain (5).

### Statistical analyses

Fisher’s exact tests were performed to compare the distribution of clades relative to patient age or disease and to compare clade distribution before and after the introduction of PCV13. A Friedman’s test with Dunn’s multiple comparison test was conducted to detect differences in the disk diffusion test result across clades and antibiotics. We also completed a Kruskal-Wallis test with Dunn’s multiple comparison test to assess differences in Pangenome size, which was based on the output from Panaroo. These statistical analyses were completed with Prism (version 10.0.03). The Bonferroni-corrected p-value threshold for the GWAS was 1.04×10^−4^. Pyseer was utilized to estimate the genetic heritability of strain country of origin (34).

## Results

The 501 serotype 3 clinical strains sampled from patients in Portugal between 1999-2020 consist of 22 different MLSTs, the most common of which were MLST180 (58%), MLST232 (18%), and MLST260 (10%) (appendix 1 p 1). MLST SLVs form four different CCs: CC180, CC232, CC260, and CC2049 (Figure 1A, appendix 1 p 1, appendix 2 pp 1). CC180 comprises 58% of the strains, with 23% belonging to CC232, 15% to CC260, and 3% CC2049 (Figure 1A, appendix 1 p 1). The GPSCs represented are GPSC12 (56% of strains), GPSC83 (37%), GPSC51 (3%), GPSC10 (0.8%), and GPSC234 (0.2%) (Figure 1A, appendix 1 p 1). GPSC12 CC180 strains included Clades Iα, Iβ, and II as defined by Azarian et al. (13) and Clades I, II, III, IV, and VI defined by Kwun et al. (16) (Figure 1A, appendix 1 p 1); subsequent references to clades refers to the latter definition unless otherwise stated.

**Figure 1.**
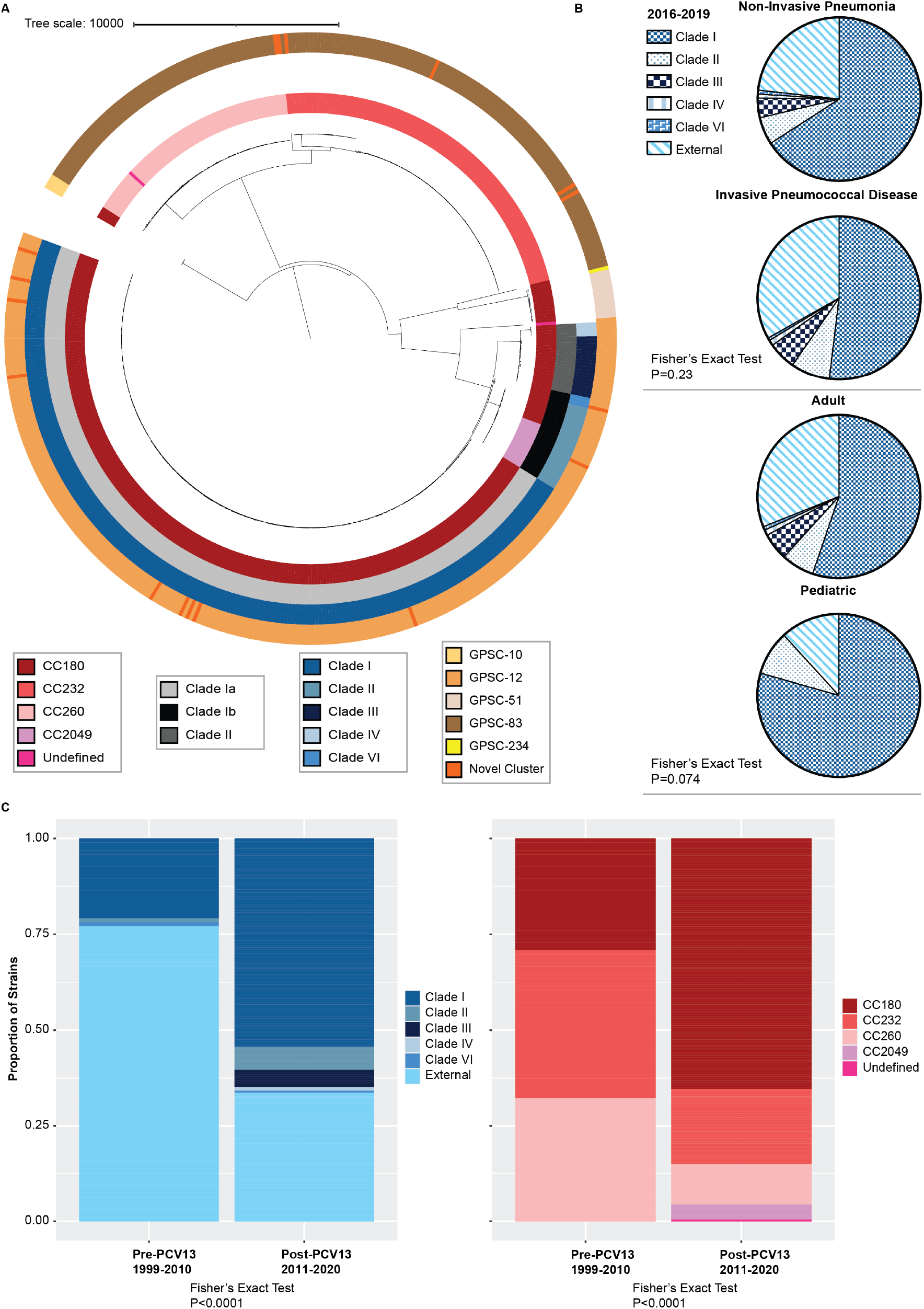
Clade dynamics of 501 serotype 3 *Streptococcus pneumoniae* (Spn) strains from Portugal. A. Maximum-likelihood phylogenetic tree of the 501 serotype 3 isolates generated from core genome single nucleotide polymorphisms. The tree scale represents substitutions per site. The legends indicate strain clonal complex (CC) in shades of red, Azarian *et al*. clade definitions in shades of grey, Kwun et al. clade definitions in shades of blue, and global pneumococcal sequencing clusters (GPSCs) orange. Subsequent references to clades refer to the Kwun et al. definition. Undefined CCs denotes either a double-locus variant or a single-locus variants of two different CCs. B. Pie charts indicating the distribution of clades by disease manifestation and age. C. Bar charts demonstrating differences in clade and CC proportions before and after the introduction of the 13-valent pneumococcal conjugate vaccine (PCV13).

We compared the distribution of clades between NIP and IPD (Fisher’s exact test, P=0.23) and between adult and pediatric cases (Fisher’s exact test, P=0.074) but did not observe a significant association between clade and disease presentation or patient age (Figure 1B, appendix 1 p 1). This analysis only considered Spn strains isolated between 2016 and 2019 because this period has the most robust representation of these features. We found a significant difference in CC proportions, and in clade proportions within CC180 before and after the introduction of PCV13 in Portugal (Fisher’s exact test, P<0.0001) (Figure 1C, appendix 1 p 1). Pre-PCV13, Clade I (Azarian et al. Clade I α) strains comprised only 21% of the dataset, but post-PCV13 Clade I became the most prevalent at 54% of all strains (Figure 1C). We did not detect Clades III and IV (Azarian et al. Clade II) pre-PCV13, but these strains only made up 5% of the sample post-PCV13 (Figure 1C). There was a concomitant decrease in strains external to the CC180 clades, from 77% to 34% (Figure 1C), with PCV13 being associated with a shift in CC prevalence (Fisher’s exact test, P<0.0001) (Figure 1C). After the introduction of PCV13 in Portugal, CC180 increased from 30% to 65%, CC232 and CC260 became less prevalent, and CC2049 became detectable (Figure 1C).

We also explored the relationship between functional genetic variation and clade background. Consistent with previous reports, the ΦOXC141 prophage is widespread amongst Clade I strains but was not detected in other lineages (Figure 2A) (13,16). Also in accordance with previous studies, antibiotic resistance genes are less prevalent in Clade I strains compared to Clades III and IV; further, PBP gene alleles are linked to strain lineage (Figure 2A) (13,35). 100% of interpretable penicillin and cefotaxime MICs were considered susceptible according to the parenteral and non-meningitis EUCAST definitions (appendix 1 p 2). Clade I MIC values did not significantly vary from that of other clades (Friedman test with Dunn’s multiple comparison test correction, P>0.31 to P>0.99) (appendix 1 p 2) (Figure 2B). Susceptibility to cotrimoxazole, erythromycin, clindamycin, tetracycline, chloramphenicol, levofloxacin, linezolid, and vancomycin ranged from 94.9%-100% of strains (appendix 1 p 3). However, Clade I strains demonstrated larger zones of inhibition across all antibiotics compared to Clade III (Friedman test with Dunn’s multiple comparison test correction, P=0.041) and Clade IV (P=0.019) but not Clade II (P>0.99), Clade VI (P=0.12), and external strains (P>0.99) (Figure 2C).

**Figure 2.**
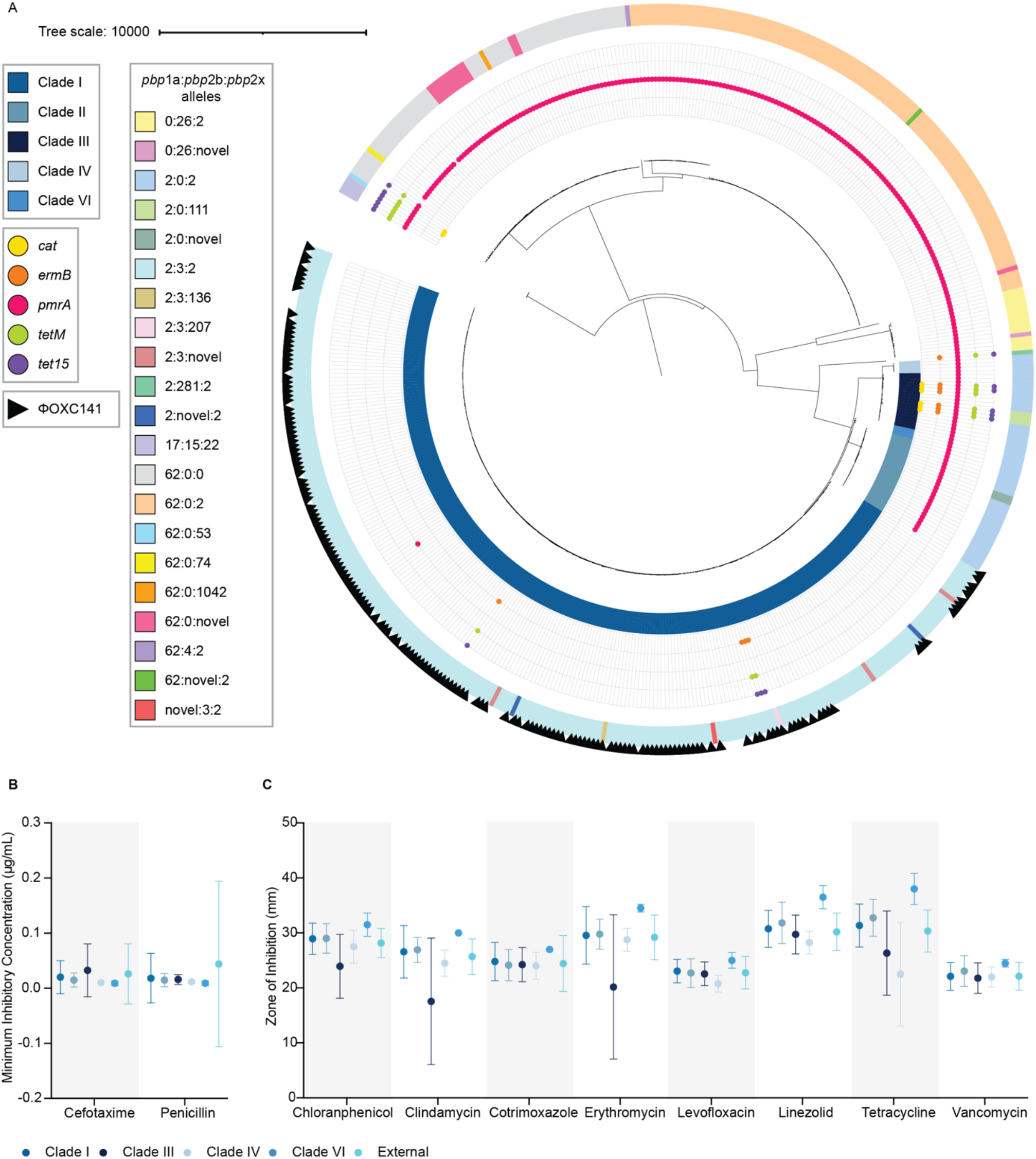
Antibiotic resistance and phage diversity in serotype 3 strains from Portugal. A. The maximum-likelihood phylogenetic tree from Figure 1A labeled to indicate the presence of various antibiotic resistance genetic determinants, penicillin binding protein gene alleles (*pbp*), and the ΦOXC141 prophage. The tree scale represents substitutions per site. 501 serotype 3 strains shown. B. Dot plots representing the distribution of minimum inhibitory concentration for cefotaxime (387 serotype 3 strains) and penicillin (401 serotype 3 strains). Dots indicate the mean and error bars indicate the standard deviation. C. Dot plots representing the distribution of the zone of inhibition (measured in in millimeters or mm) for 393 of the serotype 3 strains (385 for linezolid and vancomycin). Dots indicate the mean and error bars indicate the standard deviation.

Clade I persists as the dominant lineage in Portugal (Figure 1C) and Spain but was outcompeted by Clades III and IV in the United States, England, and Wales (5, 13-14). This could be due to genetic differences potentially modulating Clade I fitness. We generated a phylogeny comprised of 2,663 serotype 3 MLST180 strains from these regions and other countries to evaluate a connection between core genome single nucleotide polymorphisms (SNPs) and country of origin (Figure 3A, appendix 1 p 4). To do this we calculated the heritability (*h*^*2*^), which estimates how much genetic variation contributes to phenotypic variation. Using strain country of origin (Portugal, Spain, United States, and England and Wales) as the phenotype of interest, we found that *h*^*2*^ ranged from 0.88 to 1, which indicates that genotype is a meaningful indicator of strain country of origin and thus there is significant genetic relatedness among strains from a given country. Next we used the same dataset of strains to assess the correlation between strain country of origin, clade, and pangenome diversity via tsne clustering. We found that strain pangenomes largely cluster by clade, however pangenome clustering by country is also observable (Figure 3B). Consistent with this finding, we find that the pangenome of strains from Portugal have more genes than those from England and Wales (Kruskal-Wallis test with Dunn’s multiple comparison test correction, P<0.0001), the United States (P<0.0001), and other countries (P<0.0001) but not those from Spain (P>0.99) (appendix 2 pp 2). Across all countries, Clade I strains have larger pangenomes than Clade IV (Kruskal-Wallis test with Dunn’s multiple comparison test correction, P<0.0001) and Clade VI (P<0.0001), but smaller than Clade III (P=0.0002); but even within clade I strains, those from Portugal and Spain have larger pan-genomes than that of those from all other countries (Figure 3C, appendix 2 pp 2). Finally, we completed a GWAS to find genes associated with strains from Portugal and Spain. We identified 13 different genes that exceeded the Bonferroni-corrected p-value threshold of 1.04×10^−4^ (appendix 1 p 5). The top two hits are genes encoding hypothetical proteins, one whose potential function is unknown and one with a putative 3-ketoacyl-ACP reductase domain (appendix 1 p 6). The former is found in 99% of Clade I strains from Portugal and 92% of Clade I strains from Spain, while the latter is found in 67% of Clade I strains from Portugal and 90% of Clade I strains from Spain (Figure 3D). These genes are largely absent in Clade I strains from England, Wales, and the United States (Figure 3D). This trend is largely observed across Clades III, IV, and VI but country sampling from these lineages is sparse (appendix 2 pp 3). The GWAS also identified a homologue of the Spn virulence determinant *pspC* that is found in 83% of Clade I strains from Portugal but is largely absent across different clades and countries (appendix 2 pp 3).

**Figure 3.**
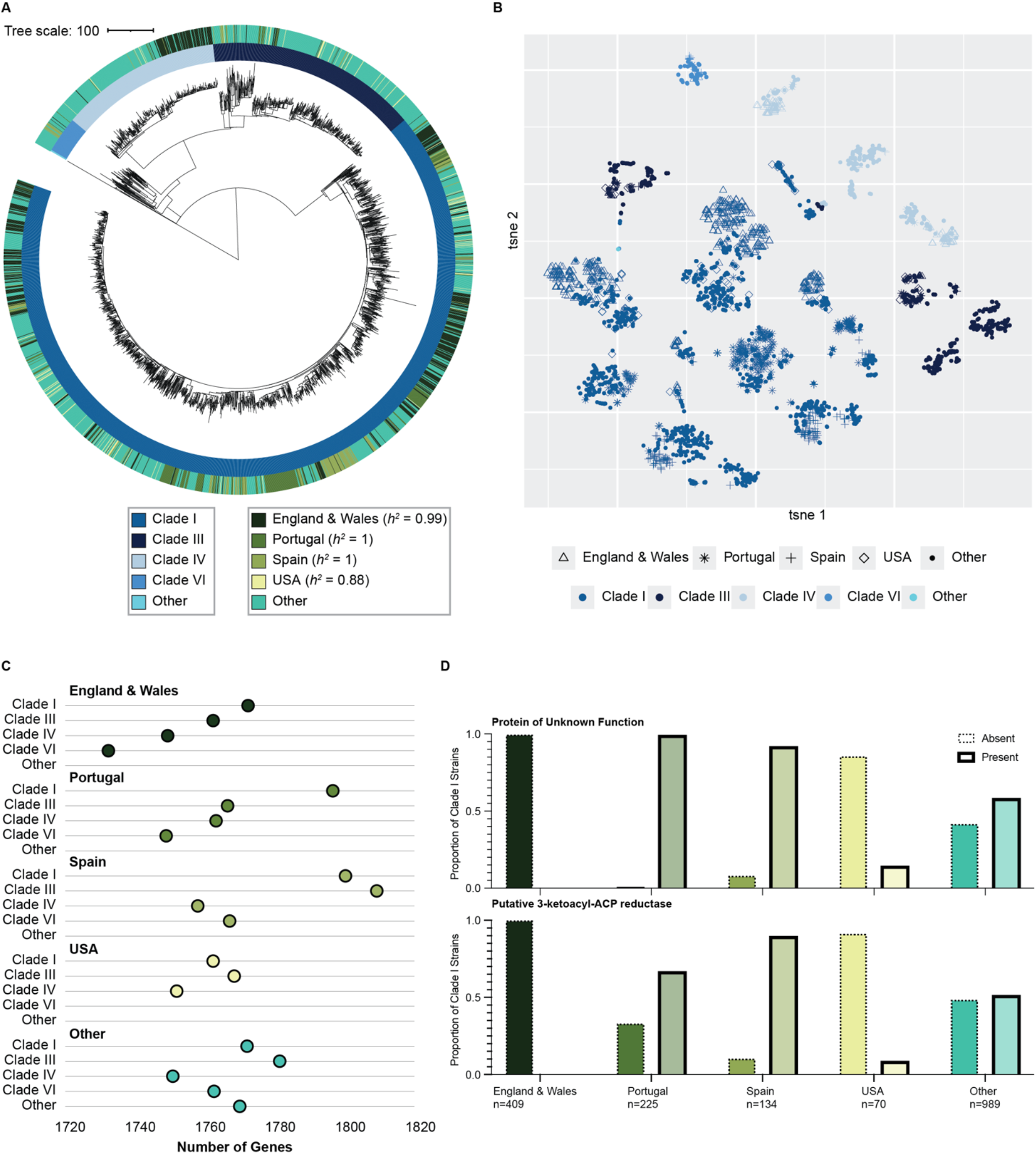
Comparative genomics of a global dataset of serotype 3 multilocus sequence type 180 (MLST180) strains. A. Maximum-likelihood phylogenetic tree of 2,663 serotype 3 MLST180 strains based on core genome single nucleotide polymorphisms. The tree scale represents substitutions per site. The heritability estimate is indicated by *h*^*2*^, the phenotype of interest is strain country of origin. B. A t-distributed stochastic neighbor embedding (tsne) plot of the 2,663 serotype 3 MLST180 strains using the pangenome content of each strain. C. Pangenome size according to the number of genes for the 2,663 serotype 3 MLST180 strains. D. Results of a genome-wide association study (GWAS) to identify genes linked to strains from Spain and Portugal amongst the 2,663 serotype 3 MLST180 strains. The top two most statistically significant hits from Clade I are shown.

## Discussion

Clade I is the most prevalent serotype 3 CC180 lineage in Portugal, even after the introduction of PCV13. This contrasts with previous observations where Clade I was supplanted by Clades III and IV in the United States, England, and Wales (13-14) but more closely resembles the clade dynamics in Spain (5). Similarly to Portugal, Spain also exhibited a post-PCV13 contraction of CC260 in favor of CC180, but while CC260 was the major lineage in Spain pre-PCV13, CC232 was the most dominant in Portugal during this time (5). Given the proximity of Portugal and Spain, the similar post-PCV13 serotype 3 CC180 lineage trends in these counties as opposed to the United States, England, and Wales may implicate a role for biogeographic context on lineage fitness and competition or the emergence of a higher fitness, but still geographically restricted, variant of Clade I. Alternatively, regional differences in clade population structure could reflect stochastic introductions of different lineages in different countries. It is important to note that our dataset comprised patients of all ages and included strains isolated from cases of both NIP and IPD whereas isolates from the United States, England, and Wales were from carriage and IPD and the study from Spain only considered IPD in adults (5,13-14). These differences in the study populations may confound comparisons across countries, although our data expanded previous observations beyond IPD by showing that there are no differences in the serotype 3 clades found in different disease presentations and patient age groups in Portugal. Nonetheless, this work provides evidence that PCV13 did not result universally in a CC180 population shift that could render the vaccine ineffective at controlling serotype 3.

We assessed the antibiotic susceptibility profile of our serotype 3 dataset from Portugal because we hypothesized that Clade I strains from this country could be more resistant than Clade I strains from the United States, England, and Wales, providing a fitness advantage that could explain why Clade I persists in Portugal but not in these regions. However, we found that Clade I isolates from Portugal are largely antibiotic-sensitive, similarly to previous observations (13-14). Along these lines, we also determined the prevalence of ΦOXC141 since one study suggested that this prophage reduced Clade I competence, which could diminish the recombination rate and ultimately hamper competitive fitness (16). We thought Clade I strains in Portugal could maintain a fitness advantage if they did not harbor ΦOXC141, as suggested by the expansion of such non-lysogenic variant of Clade I after PCV7 introduction but prior to PCV13 introduction in the UK (16). However, the Clade I strains from Portugal overwhelmingly carried the ΦOXC141 prophage.

To better understand the differences in clade dynamics in different countries, we then evaluated their genetic variation. The *h*^*2*^ estimates indicate a strong correlation between shared phylogenetic relatedness (based on core genome SNPs) and country of origin. Nonetheless, we did not observe a private subgroup of strains within Clade I circulating in Portugal and Spain, which could indicate the presence of SNPs conferring an exclusive competitive advantage. However, it is possible that differences in the pangenome could contribute to potential differences in competitive fitness. An assessment of the pangenome, revealed that Clade I isolates from Portugal and Spain have more genes than those of the United States, England, and Wales. Calvo-Silveria et al. previously reported a more open pangenome of CC180 than CC232 (5), but here we show that even within CC180, clades have variably sized pangenomes as do populations of the same clade in different geographies. It is unclear how pangenome size can impact competitive fitness, but it is possible that the repertoire of additional genes in Portugal and Spain confers an advantage to Clade I strains not available to Clade I strains in the United States, England, and Wales, which contributes to their increased prevalence. Accordingly, the GWAS identified genes that are specifically present in Clade I strains from Portugal and Spain but largely absent elsewhere: a gene encoding a protein with unknown function and a *fabG2* gene that encodes a putative 3-ketoacyl-ACP-reductase. While it is not possible to conjecture the effect of one of the hypothetical proteins, the 3-ketoacyl-ACP-reductase is involved in the elongation step of fatty acid biosynthesis. The *fabG2* gene is homologous to *fabG*, which is a member of the Spn *fab* gene cluster that directs fatty acid biosynthesis (36). Experimental studies have demonstrated that altered *fab* expression is associated with Spn carriage, biofilm formation, and CPS production (36-38). Nearly every strain in our dataset carries *fabG*, but the additional presence of *fabG2* in Clade I strains from Portugal and Spain suggest possible phenotypic effects with direct consequences on pathogenesis, which should be experimentally validated in future work. This need is underscored by a recent mouse model study demonstrating that Clade III and IV strains exhibit decreased virulence but increased carriage compared to Clade I strains, but also showing that the behavior of Clade I strains was much more variable than that of clades III and IV strains (39). This finding highlights potential genetic heterogeneity within Clade I that will have to be better defined to understand the dynamics of serotype 3 post-PCV13, as also reported here.

Overall, we observe the frequently documented trend of serotype 3 resistance to PCV13 control, but our data does not support the hypothesis suggesting this phenomenon is universally mediated by a population structure shift to the disadvantage of Clade I and favoring the emergence of Clades III and IV. This should inform and motivate efforts to identify serotype 3 determinants of vaccine escape that transcend the currently recognized clades and are representative across contexts. Moreover, mechanisms mediating serotype 3 persistence despite PCV13 use should continue to be assessed with strain datasets from diverse geographic and economic contexts to ensure the global relevance of findings, this should also include evaluations in lower-income countries and studies based in Latin America, Asia, and Africa.

## Data Availability

The whole-genome sequencing data is accessible via the National Center for Biotechnology Information Sequence Read Archive (project accession number PRJNA1152988. See appendix 1 p 1 for the accession numbers of individual strains.

## Acknowledgements

This study was funded by the National Institute of Child Health and Human Development (5T32HD040128-20 [SS, RM]). This work was also supported in part by a research grant from Investigator-Initiated Studies Program of Merck Sharp & Dohme Corp (IIS 61354) and Pfizer (CSC, JGS, JMC, MR). The opinions expressed in this paper are those of the authors and do not necessarily represent those of Merck Sharp & Dohme or Pfizer. We thank Dr. Taj Azarian for kindly providing guidance on the genomic computational analyses. We also appreciate the contribution of the patients.

## Data sharing

TThe whole-genome sequencing data is accessible via the National Center for Biotechnology Information Sequence Read Archive (project accession number PRJNA1152988). See appendix 1 p 1 for the accession numbers of individual strains.

## Appendix 2

**Supplementary Figure 1.**
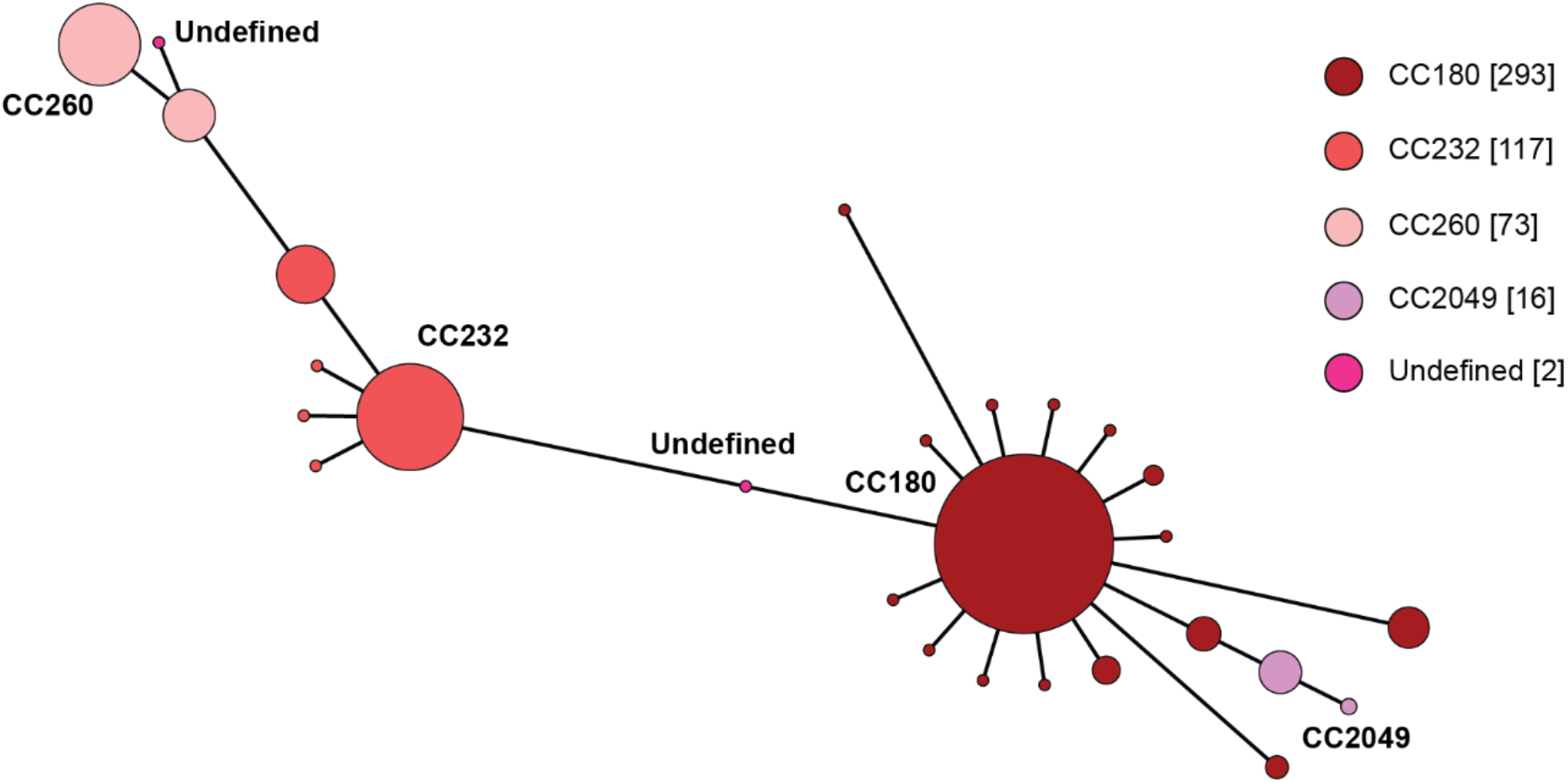
Graphic depicting the clonal complexes (CCs) represented in the 501 serotype 3 strains from Portugal. CCs are determined by single-locus variants. Each bubble indicates a different multilocus sequence type (MLST). Numbers in the brackets detail the number of strains belonging to each CC, bubble size is proportional to the number of isolates. Undefined denotes either a double-locus variant or a single-locus variants of two different CCs.

**Supplementary Figure 2.**
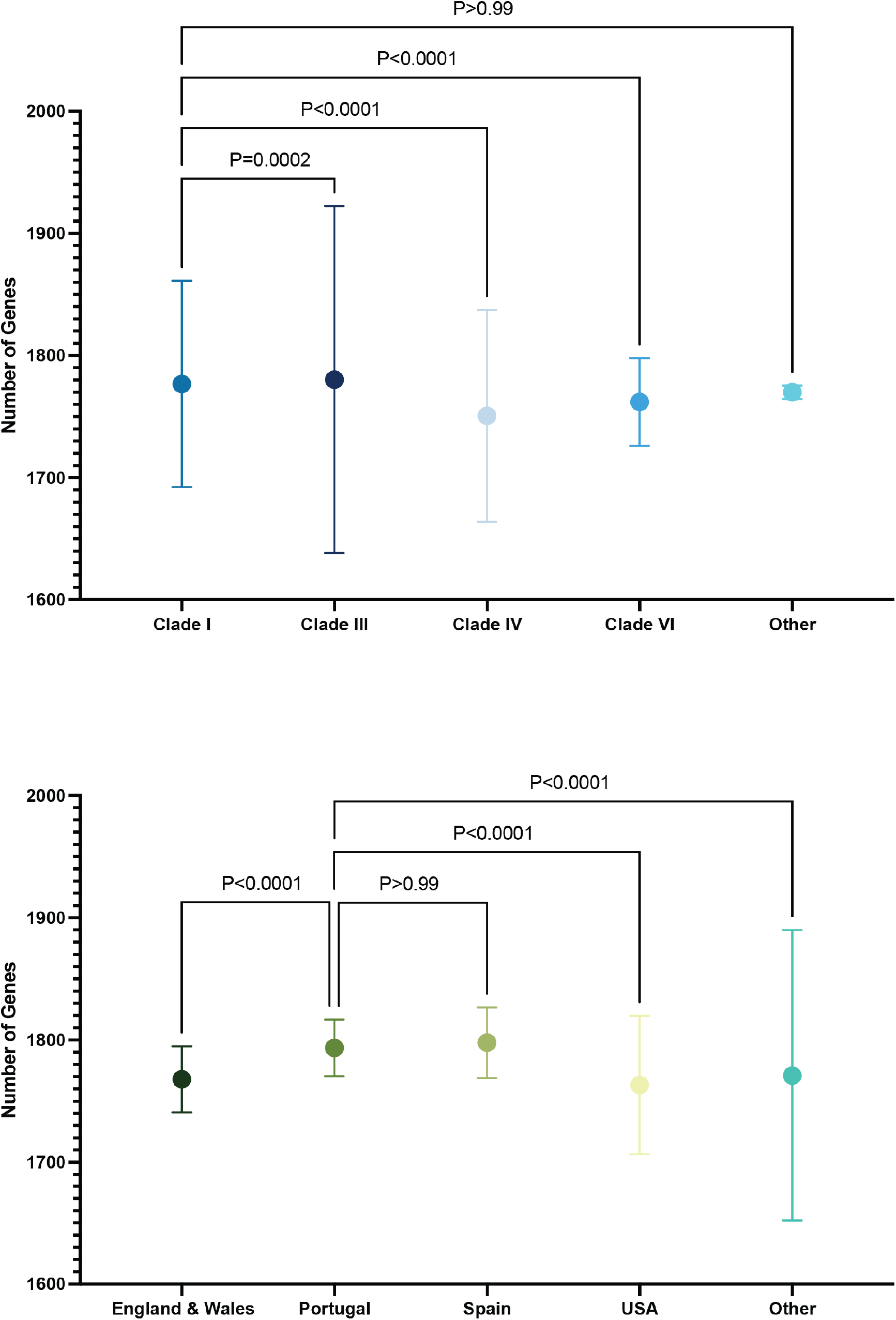
Dot plots indicate the distribution of the number of genes across the 2,663 serotype 3 MLST180 strains. The p-values denote the results of a Kruskal-Wallis test with Dunn’s multiple comparison test correction.

**Supplementary Figure 3.**
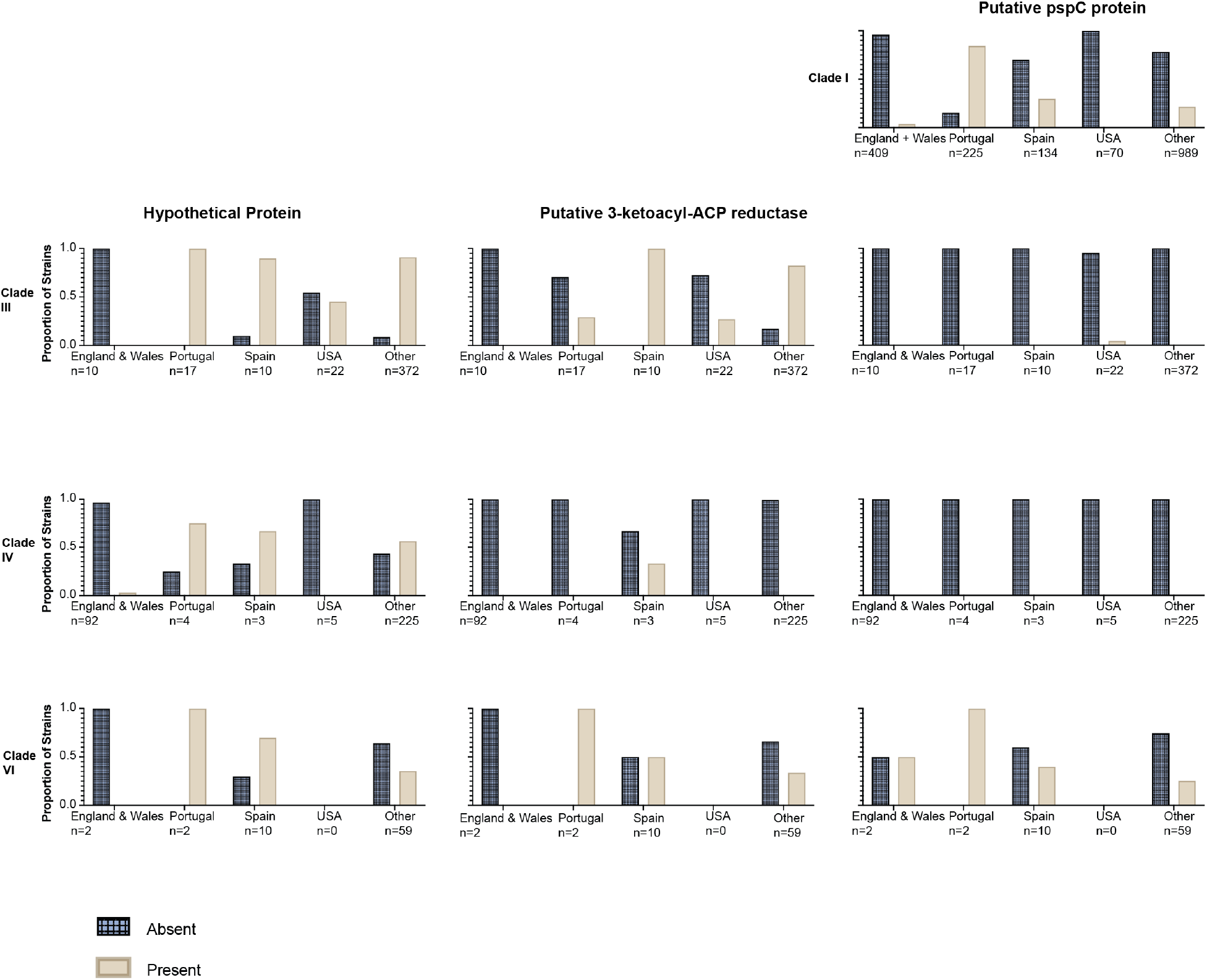
Bar plots representing the results of a genome-wide association study (GWAS) to identify genes linked to strains from Spain and Portugal amongst the 2,663 serotype 3 MLST180 strains.

